# SARS-CoV-2 specific T cell and humoral immune responses upon vaccination with BNT162b2: A 9 months longitudinal study

**DOI:** 10.1101/2021.11.06.21265632

**Authors:** Junko S. Takeuchi, Ami Fukunaga, Shohei Yamamoto, Akihito Tanaka, Kouki Matsuda, Moto Kimura, Azusa Kamikawa, Yumiko Kito, Kenji Maeda, Gohzoh Ueda, Tetsuya Mizoue, Mugen Ujiie, Hiroaki Mitsuya, Norio Ohmagari, Wataru Sugiura

## Abstract

**Introduction:** The humoral and cellular immune responses against severe acute respiratory syndrome coronavirus 2 (SARS-CoV-2) upon coronavirus disease 2019 (COVID-19) vaccination remain to be clarified. Hence, we aimed to investigate the long-term chronological changes in SARS-CoV-2 specific IgG antibody, neutralizing antibody, and T cell responses during and after receiving the BNT162b2 vaccine.

**Methods:** We performed serological, neutralization, and T cell assays among 100 hospital workers aged 22-73 years who received the vaccine. We conducted seven surveys up to eight months after the second vaccination dose.

**Results:** SARS-CoV-2 spike protein-specific IgG (IgG-S) titers and T cell responses increased significantly following the first vaccination dose. The highest titers were observed on day 29 and decreased gradually until the end of the follow-up period. There was no correlation between IgG-S and T cell responses. Notably, T cell responses were detected on day 15, earlier than the onset of neutralizing activity.

**Conclusions:** This study demonstrated that both IgG-S and T cell responses were detected before acquiring sufficient levels of SARS-CoV-2 neutralizing antibodies. These immune responses are sustained for approximately six–ten weeks but not for seven months or later following the second vaccination, indicating the need for the booster dose (i.e., third vaccination).

## INTRODUCTION

The coronavirus disease 2019 (COVID-19), caused by severe acute respiratory syndrome coronavirus 2 (SARS-CoV-2), has imposed a tremendous burden on global morbidity and mortality since December 2019 ^1^. In response to the COVID-19 pandemic, multiple vaccines have been rapidly developed and tested for their efficacy and safety. BNT162b2, an mRNA-based SARS-CoV-2 vaccine (Pfizer-BioNTech), was the first COVID-19 vaccine listed for emergency use and COVAX roll-out by the World Health Organization, with a 95% efficacy in clinical trials ^2^. To examine the effectiveness of the vaccine, a growing body of studies has investigated SARS-CoV-2 specific humoral and cellular responses following vaccination. Studies have shown that a two-dose regimen of BNT162b2 induces SARS-CoV-2 specific humoral and cellular responses, such as immunoglobulin G (IgG) antibodies ^3-5^, neutralizing antibodies ^6-8^, as well as CD4^+^ and CD8^+^ T cells ^6,7,9^.

However, some important issues remain to be addressed. First, little is known about neutralizing antibodies and cellular immune responses against SARS-CoV-2 following COVID-19 vaccination compared to the evidence on IgG antibodies. Second, epidemiological evidence on temporal changes in humoral and cellular responses and their associated factors, and use of longitudinal data with repeated measures, is limited. Third, studies on the correlation between humoral and cellular responses are scarce. Investigation of humoral and cellular immune responses during and after the COVID-19 vaccination regimen is required to understand its effectiveness, the duration of effectiveness, and the differences in such responses across varying background factors.

Hence, this longitudinal study aimed to investigate chronological changes in IgG antibody, neutralizing antibody, and T cell responses, along with their associated factors, during and after the BNT162b2 vaccination regimen, in hospital workers of a national medical institution in Japan.

## METHODS

### Patient Consent Statement

Written informed consent was obtained from all the participants before the baseline survey. The study protocol was approved by the Ethics Committee of NCGM, Japan (approval number: NCGM-A-004175-00).

### Study Design and Participants

We performed a longitudinal study among the staff of the National Center for Global Health and Medicine (NCGM), a national medical institution designated for research and treatment of infectious diseases, between March and December 2021, after the start of the vaccination program. We recruited 100 volunteers via e-mail from NCGM staff aged 20 years or above who were scheduled to receive the first COVID-19 vaccination dose on the dates of the baseline survey. None of the participants were on immunosuppressive medication or had a history of COVID-19. All participants received the mRNA-based SARS-CoV-2 vaccine BNT162b2 (Pfizer-BioNTech) according to the standard protocol (two doses of 30 µg administered three weeks apart) ^6,10^. A total of seven surveys were conducted on the following schedule: day 1 (immediately after the first dose), day 15, day 29 (seven days after the second dose), day 61, day 82–96 (at the time of periodic health checkup), day 224–232, and day 263 (∼ eight months after the second dose).

### Cells and Viruses

VeroE6TMPRSS2 cells ^11^ were obtained from the Japanese Collection of Research Bioresources (JCRB) Cell Bank (Osaka, Japan). VeroE6TMPRSS2 cells were maintained in DMEM supplemented with 10% FCS, 100 μg/mL penicillin, 100 μg/mL streptomycin, and 1 mg/mL G418. The SARS-CoV-2 NCGM-05-2N strain (SARS-CoV-205-2N) was isolated from the nasopharyngeal swab of a patient with COVID-19 admitted to the NCGM hospital ^8,12^.

### Serological Assays

We performed three serological assays to detect the IgM and IgG antibodies against SARS-CoV-2 spike protein (IgM-S and IgG-S, respectively) and IgG antibodies against SARS-CoV-2 nucleocapsid protein (IgG-N). The presence of IgG-N antibodies can indicate SARS-CoV-2 infection prior to the study and during follow-up, regardless of the vaccination status. In contrast, the presence of IgM-S and IgG-S indicates the previous infection and/or humoral immunity following vaccination, because BNT162b2 is constructed to express the full-length spike protein.

To detect IgG-S, the AdviseDx SARS-CoV-2 IgG II assay was performed using the Abbott ARCHITECT® following the manufacturer’s protocol. The assay detects the IgG antibodies against the receptor-binding domain (RBD) of the S1 subunit of the SARS-CoV-2 spike protein using chemiluminescent microparticle immunoassay (CMIA). The resulting chemiluminescence in relative light units (RLU) indicates the strength of the response, which in turn reflects the quantity of IgG-S present. We considered 50 AU/mL and above as the cutoff for seropositivity, as recommended by the vendor. We also tested 4,160 AU/mL as the cutoff, given that this threshold may correspond to a 95% probability of sufficient neutralizing activity ^13^. This quantitative IgG-S test could evaluate an individual’s humoral response to vaccines.

To detect IgM-S, the AdviseDx SARS-CoV-2 IgM assay, based on semi-quantitative CMIA, was performed using the Abbott ARCHITECT® according to the manufacturer’s instructions. We defined the seropositive cutoff as 1.0 (index value) and above, according to the vendor’s recommendation.

For the IgG-N assay, the Abbott ARCHITECT® SARS-CoV-2 anti-N IgG assay based on semi-quantitative CMIA was performed. We considered the index values of 1.40 and above as seropositive cutoff according to the manufacturer’s instructions.

### Neutralization Assay

An enzyme-linked immunosorbent assay (ELISA)-based semi-quantitative neutralization assay was conducted at three points of the survey (day 15, 29, and 61). It was performed at a single serum dilution according to the manufacturer’s instructions (SARS-CoV-2-NeutraLISA, Euroimmun, Germany). Antibody-mediated inhibition of soluble human receptor protein (angiotensin-converting enzyme 2, hACE2) binding to the RBD of the viral S1 region was tested. The surrogate neutralizing capacity was calculated as the percentage of inhibition (% IH): IH = 100% - (extinction of the respective sample × 100% / extinction of the blank). An IH<20% was considered as negative, IH ≥20% to <35% intermediate, and IH ≥35% positive. Each serum sample was tested in duplicates.

The *in vitro* viral experiment-based neutralizing activity was determined by quantifying the serum-mediated suppression of the cytopathic effect (CPE) of the SARS-CoV-2 strain in VeroE6TMPRSS2 cells, as previously described ^8^, but with minor modifications. In brief, each serum was four-fold serially diluted in the culture medium. The diluted sera were incubated with 100 TCID50 (50% tissue culture infectious dose) of viruses at 37 °C for 20 minutes. Then, the serum-virus mixtures were inoculated into VeroE6TMPRSS2 cells (1.0 × 10^4^/well) in 96-well plates. After culturing the cells for three days, the CPE levels observed in SARS-CoV-2-exposed cells were determined by the WST-8 assay using the Cell Counting Kit-8 (Dojindo, Kumamoto, Japan). The serum dilution that resulted in 50% inhibition of CPE was defined as a 50% neutralization titer (NT50). Each serum sample was tested in duplicates.

### IFN-γ Whole Blood Assay

SARS-CoV-2 specific CD4^+^ and CD8^+^ T cell responses were investigated at six points (day 1, 15, 29, 61, 224–232, and 263). We quantified T cell-produced interferon-gamma (IFN-γ) in response to SARS-CoV-2 spike peptides using QuantiFERON SARS-CoV-2 RUO (QIAGEN, Germany) according to the manufacturer’s instructions. Briefly, 5 mL of whole blood specimens were collected in a collection tube containing sodium heparin (VP-H050K, Terumo, Japan) at six time points during the vaccination regimen. The collected blood samples were stored at room temperature for no more than 2–3 hours, and 1 mL of the sample was transferred to each QuantiFERON blood collection tube: Nil (negative control), mitogen (positive control), Ag1 and Ag2 (spike protein antigens). Ag1 induces a CD4^+^ T cell response, and Ag2 stimulates both CD4+ and CD8+ T cell responses. The four tubes were inverted to coat the sides of the tubes and placed in a 37 °C incubator for 16–24 hours. Subsequently, the plasma samples were harvested following centrifugation, stored at -80 °C, and tested for the release of IFN-γ from stimulated T cells by QuantiFERON ELISA with microplate reader SpectraMax iD5 (Molecular Devices, USA). The quantitative IFN-γ concentration (IU/mL) was analyzed using the QuantiFERON R&D Analysis Software (version 5.3.0). Greater than or equal to 0.20 IU/mL IFN-γ concentration was used to define positive T cell responses, as previously reported ^14^.

### Statistical Analysis

To examine the difference in changes of IgG-S titers across background factors (age, sex, and body mass index [BMI]), we fitted the mixed model of repeated measures with an unstructured covariance matrix. Background factors were treated as categorical variables in the models: age (<40 or ≥40 years), sex (men or women), and BMI (<25 or ≥25 kg/m^2^). Log-transformed IgG-S titers were used as the dependent variable. We set each background factor, time, and interaction terms as fixed factors, and individual identifiers and time as random factors in the model. In all models, we adjusted for age and sex as covariates. We estimated the mean log-transformed IgG-S titers with 95% confidence intervals (CIs); then, we back-transformed them and presented them as geometric means titers. To compare the mean differences of IgG-S titers between background factors at each time point, we used the Wald test. As three participants received their second vaccination on an irregular schedule (more than four weeks after the first dose), we treated their antibody titers data from day 29 to day 263 as missing values. We repeated these analyses for spike-specific T cell responses.

To examine the correlation between IgG-S titers and spike-specific T cell responses at each time point (day 1, 15, 29, 61, 224–232, and 263), we conducted Spearman’s rank correlation test.

Statistical analyses were conducted using STATA version 17.0 and GraphPad Prism 9.3.0.

## RESULTS

### Study participants

Table 1 presents participants’ background information. The participants included 32 men and 68 women with a mean age (standard deviation) of 42.7 (11.7) years (range: 22–73 years). Major occupations included administrative staff (19%), nurses (17%), doctors (7%), allied healthcare professionals (4%), and other occupations (e.g., researchers and research assistants) (53%).

**Table 1.**
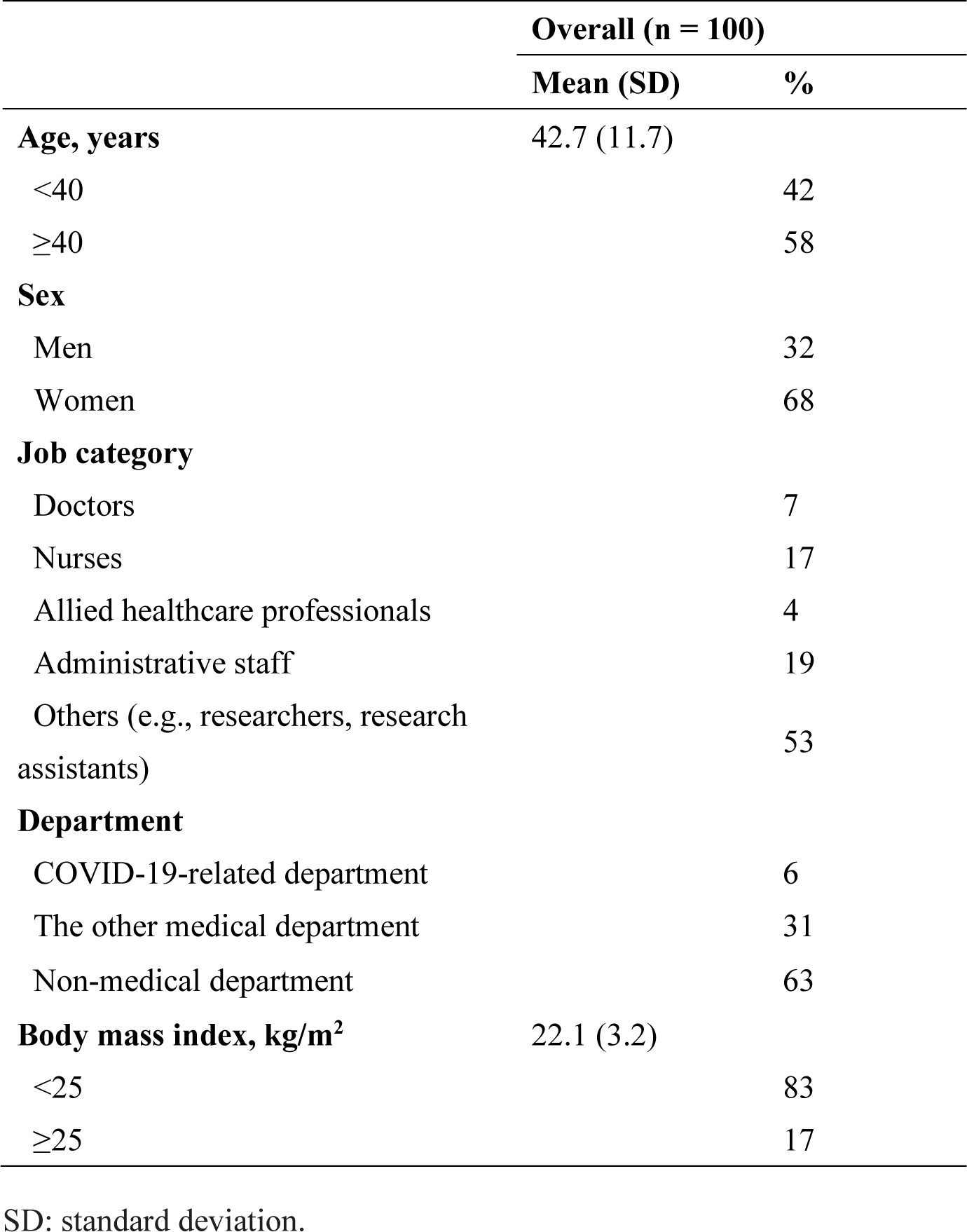
Participants’ characteristics (n = 100)

### Antibody Titers and Neutralizing Activity

Table 2 and Figures 1–2 show the humoral and cellular responses during and after the vaccination regimen. All participants were seronegative for IgG-S, IgG-N, and IgM-S on day 1 (Figure 1A–C). These observations were consistent with the fact that none of the participants had a history of COVID-19. IgG-S titers were significantly higher than the baseline value in the later time points (Table 2). We observed the highest IgG-S titers on day 29 (seven days after the second dose), with a median of 20,582 AU/mL (interquartile range [IQR] 11,372–29,747), followed by a time-dependent decrease: median of 9,649 (IQR 5,168–13,143) on day 61, 5,419 (IQR 2,854–8,209) on day 82–96, 780 (IQR 439–1,345) on day 224–232, and 784 (IQR 301– 1,038) on day 263. For IgG-S, 99% (99/100) of the participants were seropositive on day 15 following the first dose, and all participants were seropositive on day 29 (seven days after the second dose) and remained positive during follow-up until day 263. Regarding the surrogate measures of neutralizing activity defined by the IgG-S titers (cutoff of 4,160 AU/mL), no participants were seropositive on day 15, while 96% became positive on day 29; this decreased to 81% and 58% on day 61 and day 82–96, respectively (Table 2). With longer follow-up, all participants became seronegative on day 224–232 and day 263 (< 4,160 AU/mL). We also tested neutralizing activity using the ELISA-based semi-quantitative neutralization assay, which showed that 37% of the participants had sufficient neutralizing activity on day 15, while all participants had sufficient levels of neutralizing activity on day 29 and 61 (Table 2 and Figure 1D).

**Table 2.**
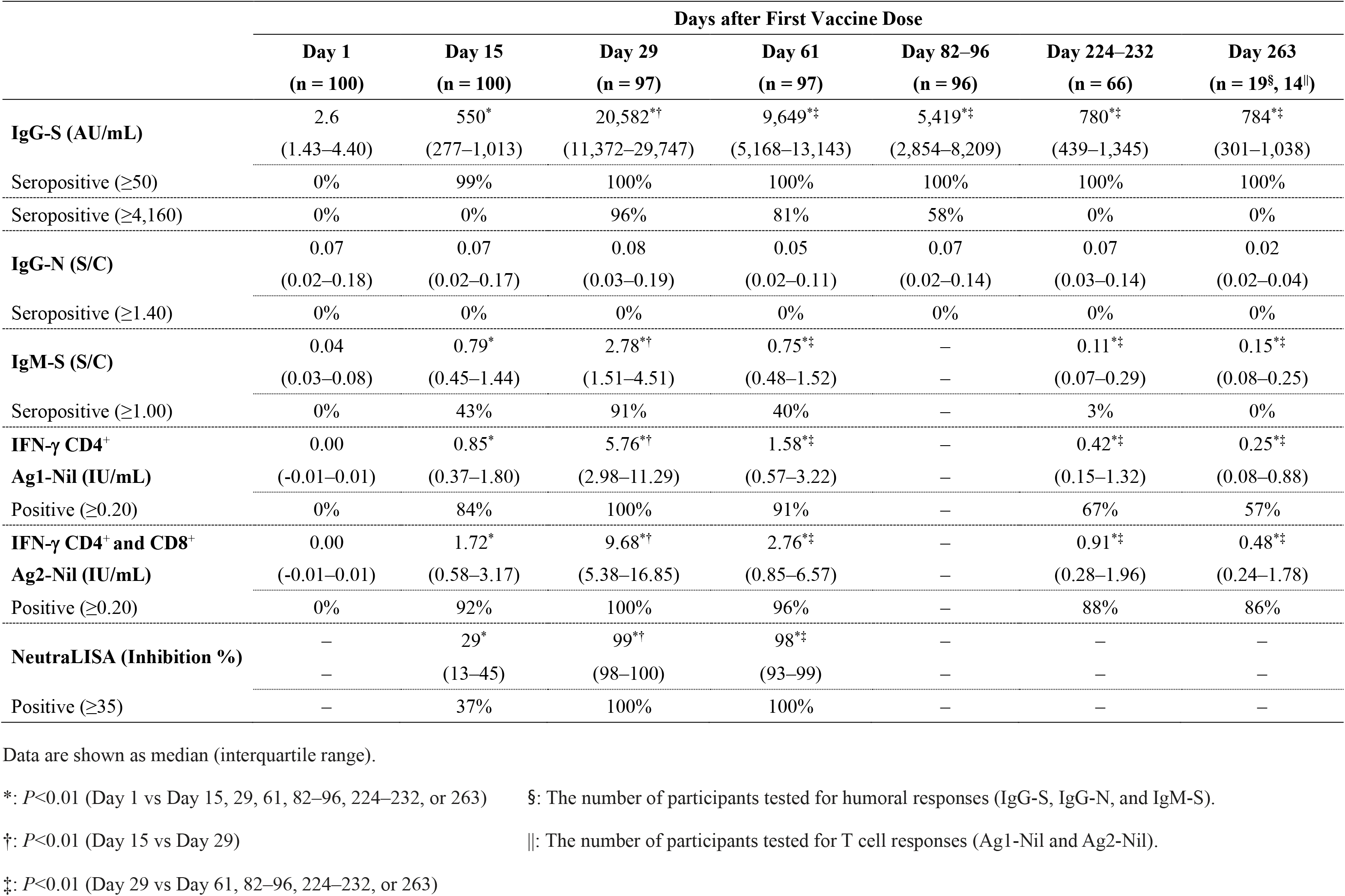
Humoral and cellular immune responses during and after BNT162b2 mRNA-based SARS-CoV-2 vaccination regimen

**Figure 1.**
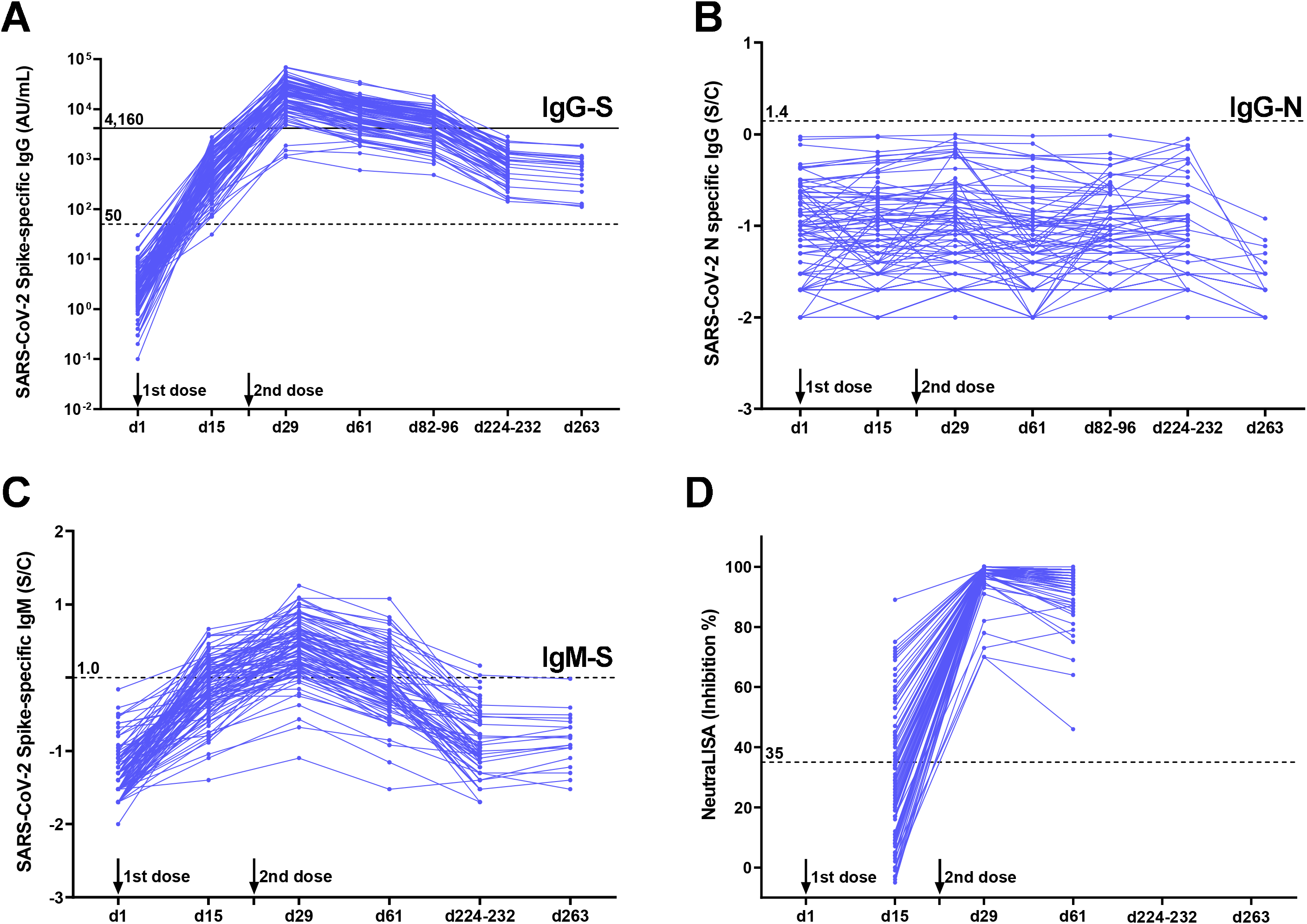
Humoral responses during and after BNT162b2 mRNA-based SARS-CoV-2 vaccination regimen. (A-C) Humoral responses in serum on day 1 (immediately after the first dose), day 15, day 29 (seven days after the second dose), day 61, day 82–96, day 224– 232, and day 263. IgG antibodies against SARS-CoV-2 spike protein (A), nucleocapsid (B), and IgM against spike protein (C) were measured using chemiluminescent microparticle immunoassay (CMIA). An ELISA-based semi-quantitative neutralization assay was also performed (D). Dashed lines indicate the cutoff for each assay. All data were represented as line graphs using GraphPad Prism 9.3.0. Black line indicates 4,160 AU/mL as the cutoff, given that this threshold may correspond to a 95% probability of sufficient neutralizing activity.

There was a significant association of younger age (<40 years) with higher IgG-S titers throughout the five survey time points (*P*<0.01) (Figure 4A and Supplementary Table S1). Although we did not observe a statistically significant difference in IgG-S titers with respect to sex or BMI, females or individuals with normal BMI tended to have higher IgG-S titers than males or those with BMI ≥25 kg/m^2^ throughout the surveys (Figure 4B–C and Supplementary Table S1).

### Vaccine-Induced T cell Responses

None of the participants had T cell responses against SARS-CoV-2 antigens on day 1, indicating that none of them were infected with SARS-CoV-2 prior to the study. Following the first vaccination dose, T cell responses increased (0.85 and 1.72 IU/mL for Ag1-Nil and Ag2-Nil, respectively) and peaked after the boost on day 29 (5.76 and 9.68 IU/mL for Ag1-Nil and Ag2-Nil, respectively), but decreased to day 61 (1.58 and 2.76 IU/mL for Ag1-Nil and Ag2-Nil, respectively), day 224–232 (0.42 and 0.91 IU/mL for Ag1-Nil and Ag2-Nil, respectively), and day 263 (0.25 and 0.48 IU/mL for Ag1-Nil and Ag2-Nil, respectively) (Table 2 and Figure 2). Overall, higher values for T cell response were obtained for Ag2-Nil (CD4^+^ and CD8^+^) than Ag1-Nil (CD4^+^), indicating that both CD4^+^ and CD8^+^ T cells serve as cellular immunity induced by the COVID-19 vaccination.

**Figure 2.**
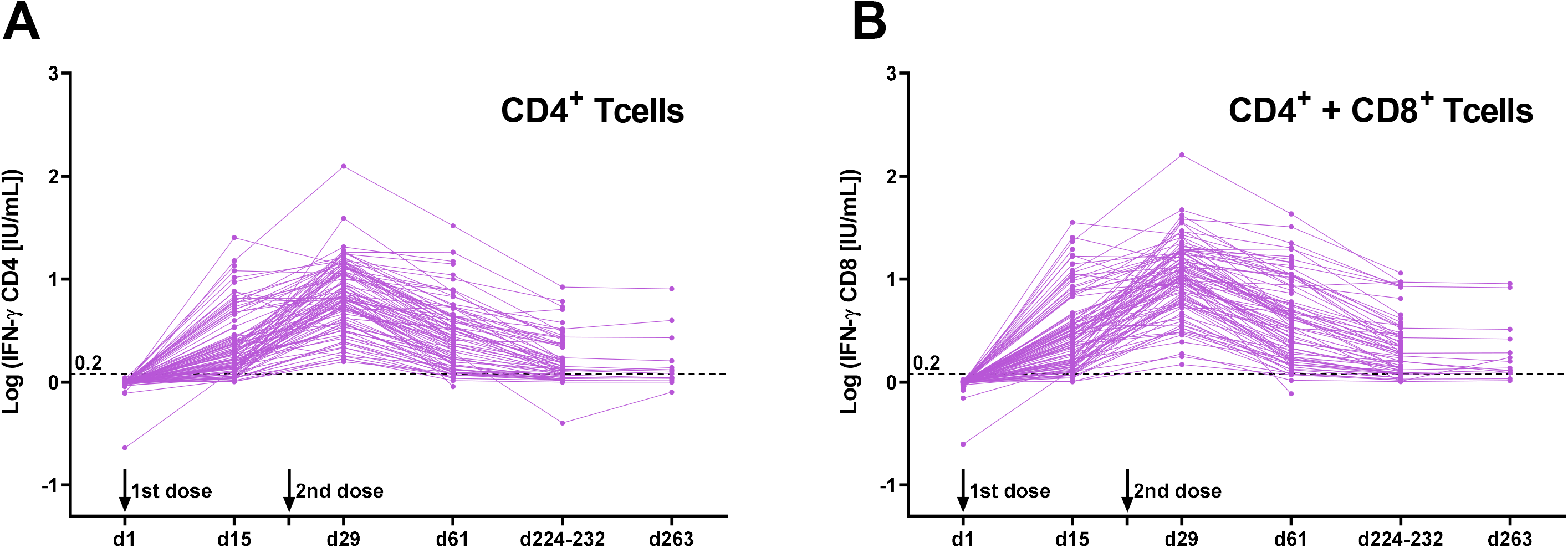
SARS-CoV-2 spike specific T cell response during and after BNT162b2 mRNA-based SARS-CoV-2 vaccination regimen. The release of IFN-γ from stimulated CD4^+^ T cells (A) and both CD4^+^ and CD8^+^ T cells (B) with SARS-CoV-2 spike peptides using QuantiFERON SARS-CoV-2 RUO. All data were represented as line graphs using GraphPad Prism 9.3.0. Dashed lines indicate the cutoff for the assay.

While analyzing the association between background factors (age, sex, and BMI) and T cell responses, we did not observe a statistically significant difference in IFN-γ produced by T cells by age or BMI at all time points (data not shown). While men were significantly associated with a higher IFN-γ production by CD4^+^ T cells on day 29 (*P* <0.05), this association was not observed when one high outlier was excluded (Supplementary Figure S1).

Further, we found no correlations between IgG-S and IFN-γ production in CD4^+^ (Figure 3A) and CD4^+^ + CD8^+^ T cells (Figure 3B) throughout the surveys, except on day 15, and significantly weak correlations were observed on day 15 (Spearman rank coefficient [rho] = 0.83; p <0.01 and rho = 0.29; p <0.01 for CD4^+^ and CD4^+^ + CD8^+^, respectively).

**Figure 3.**
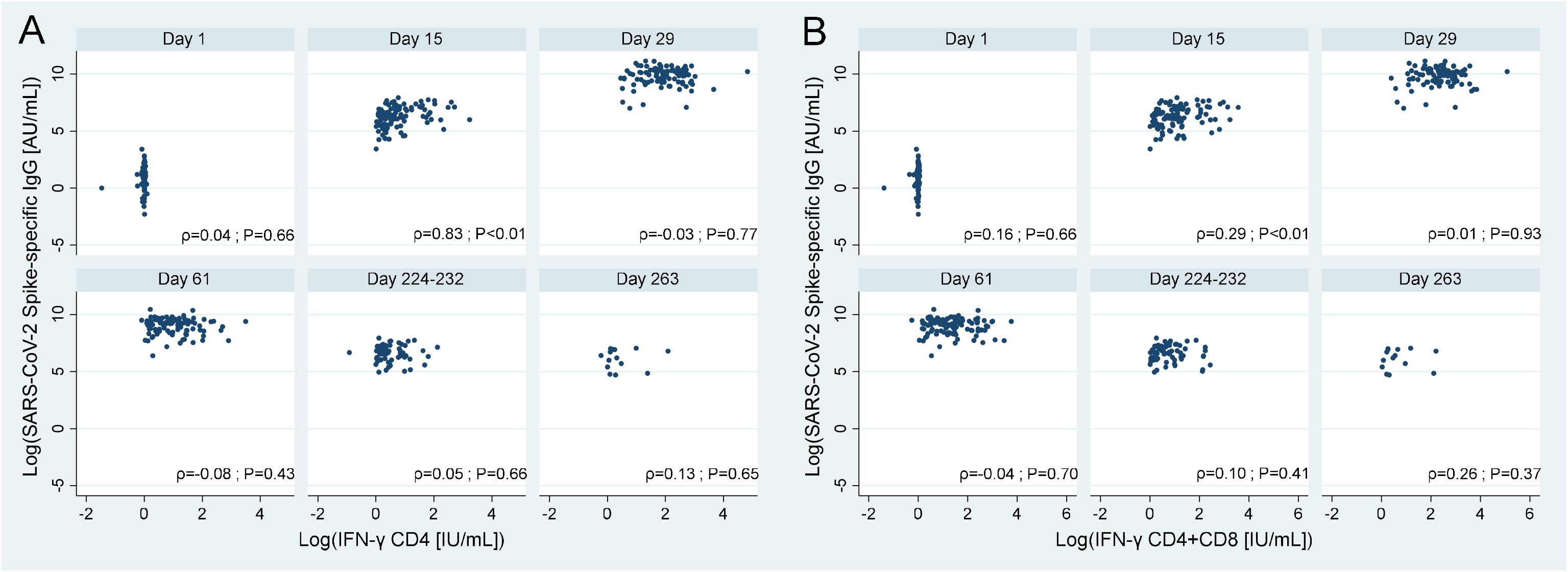
Correlation between SARS-CoV-2 spike specific IgG titers and T cell responses at each time point. Spearman’s rank correlation test was conducted using STATA version 17.0. (A) Correlation between IgG-S titer and spike-specific CD4^+^ T-cell responses. (B) Correlation between IgG-S titer and spike-specific CD4^+^ and CD8^+^ T cell responses.

**Figure 4.**
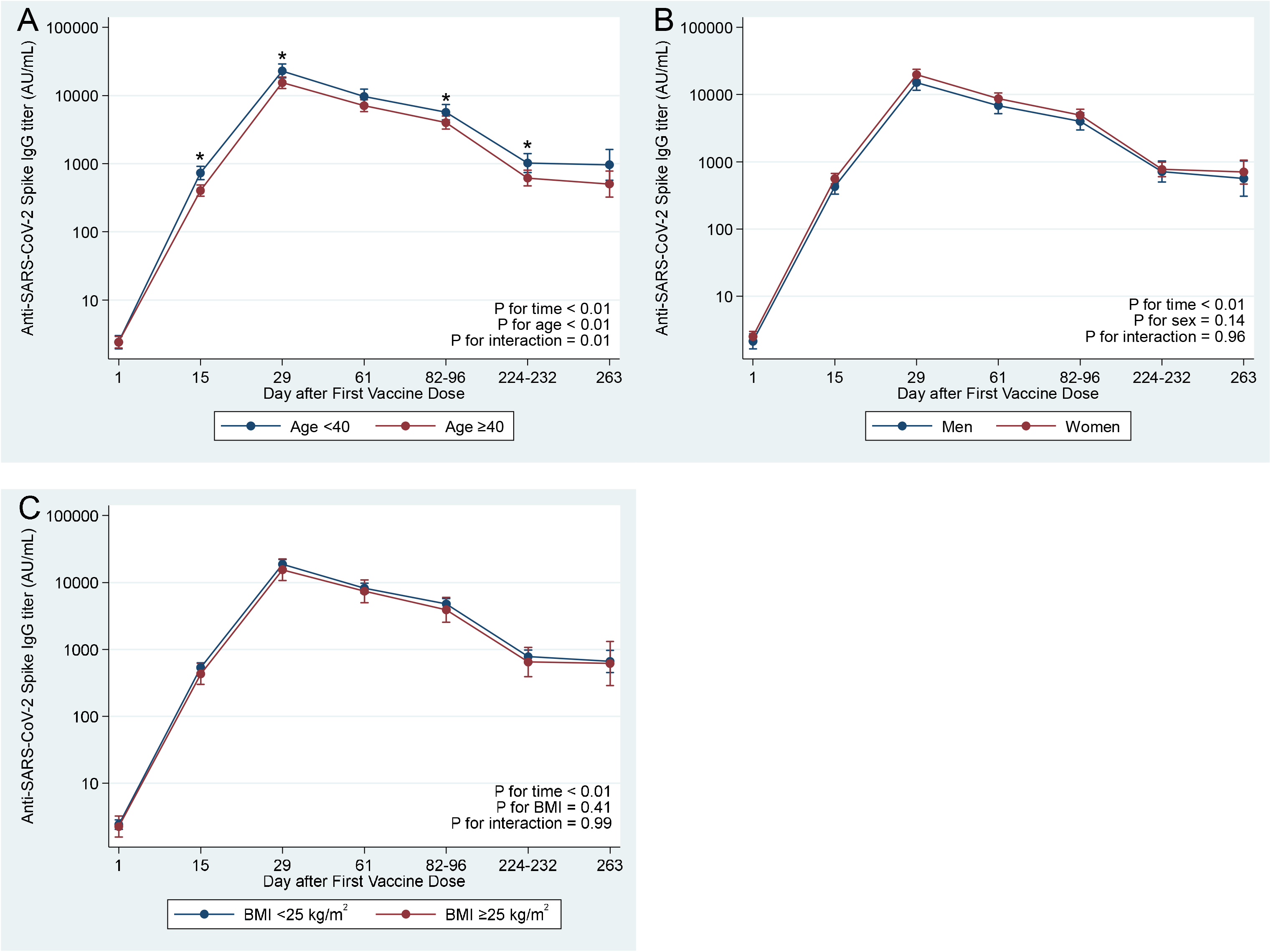
SARS-CoV-2 spike specific IgG titers by background factors. Wald test was used to compare the mean differences of IgG-S titers between background factors at each time point using STATA version 17.0. (A) SARS-CoV-2 spike specific IgG titers by age (<40 or ≥40 years) (B) SARS-CoV-2 spike specific IgG titers by sex (men or women) (C) SARS-CoV-2 spike specific IgG titers by BMI (<25 or ≥25 kg/m^2^).

## DISCUSSION

In this study, we documented the IgG-S antibody and T cell responses on day 15 following the first dose of the BNT162b2 vaccine. We found that both IgG-S antibody and T cell responses peaked seven days after the second dose (day 29) and gradually declined until the end of the follow-up period. Sufficient levels of the IgG-S antibody were maintained for over six to ten weeks following the second dose of BNT162b2 vaccination, but all participants became seronegative at seven months or later after the second dose when 4,160 AU/mL was used as the cutoff. It is noteworthy that T cell responses were detected earlier than the neutralizing activity. However, we did not observe any statistically significant correlation between humoral and cellular immune responses throughout the survey period.

We observed that IgG-S titers on day 15 and day 29 were 211-fold and 7,916-fold higher, respectively, than that of day 1, and it decreased by 53.1%, 73.7%, and 96.2% on day 61 (39 days after the second dose), day 82–96 (60–74 days after the second dose), and day 263 (eight months after the second dose), respectively, compared to the peak on day 29. Similar to our findings on chronological changes in IgG-S titers, a multicenter prospective study among health care professionals at multiple time points (day 0, 14, 28, 42, 56, and 90) showed that the antibody response (measured as IgG-S) was detected on day 14 following the first dose and reached a peak between day 14 and 28 (the second dose was administered on day 21). Further, the response decreased by 43% on day 90 compared to the highest antibody response ^3^. As for longer follow-up time period, another study in a large cohort of 4,868 participants showed that IgG-S titers decreased by 94.5% and waned at six months after the second dose compared to the peak ^15^, which is consistent with our finding.

In contrast to the IgG-S antibody response, the majority of participants did not attain sufficient levels of neutralizing antibodies against SARS-CoV-2 on day 15, as assessed by ELISA-based neutralization assay (37%) and IgG-S titers with a cutoff of 4,160 AU/mL (0%). However, nearly 100% of participants were positive for neutralizing activity in both assays on day 29. These results suggest that individuals might attain sufficient neutralizing antibodies against SARS-CoV-2 seven days after the second dose of the BNT162b2 vaccine. Further investigations should be tested in neutralizing activity for a longer follow-up time period, given that the proportion of those with sufficient neutralizing antibodies declined after the peak when it was assessed with IgG-S titers.

In the analysis of investigating the changes of IgG-S titers across background factors, younger age was significantly associated with higher IgG-S titers throughout the follow-up period. This finding accords with previous studies that showed the association between age and antibody responses ^5,16,17^. It has been suggested that age is an important contributing factor for vaccine-induced humoral immune responses. Similarly, our findings and previous evidence suggest that older individuals are vulnerable to lower IgG-S titers throughout and after COVID-19 vaccination compared to younger individuals.

The present study showed that T cell responses were detected on day 15 following the first dose of the vaccination, similar to the antibody responses. This early induction of T cell responses agrees with the findings of a cohort study of 20 Singapore healthcare workers who received the BNT162b2 vaccine ^7^, and a study of individuals who received the Moderna mRNA-1273 vaccine ^14^. Notably, we observed that T cell responses were detectable earlier than the neutralizing antibodies, consistent with a previous report ^7^. A clinical trial evaluating the efficacy of BNT162b2 indicated that the vaccine might prevent the onset of COVID-19 approximately 12 days after the first dose ^10^. These observations suggest that early T cell responses, detected on day 15 and earlier than neutralizing antibodies, may contribute to the vaccine’s efficacy. Interestingly, early T cell responses have also been reported in COVID-19 cases. A review paper by Bertoletti *et al*. suggested that T cells are detected within seven–ten days after infection ^18^. Moreover, early induction of IFN-γ-producing SARS-CoV-2-specific T cells was observed in patients with mild symptoms ^19^. Rapid and functional induction of T cell immune responses may play a critical role in both COVID-19 cases and vaccinated individuals.

However, we did not find any evidence of an association between background factors (age, sex, and BMI) and T cell responses. A recent study reported that the production of IFN-γ from SARS-CoV-2 spike-specific T cells was lower in older vaccinated participants (≥ 80 years old) ^20^. In our study, all participants were younger than 80 years old (the maximum age was 73 years), which might explain the null finding on the association between age and T cell responses.

We did not observe any statistically significant correlation between IgG-S and IFN-γ production in CD4^+^ and CD4^+^ + CD8^+^ T cells. Some participants had higher IFN-γ values with low or moderate IgG-S levels and vice versa. However, the correlation between SARS-CoV-2 spike-specific humoral and cellular immune responses remains controversial ^7,21^. This discrepancy can be explained by the pre-existence of cross-reactive memory T cells against SARS-CoV-2. Interestingly, previous studies have reported that long-lasting memory T cells, induced by common cold coronaviruses, can display robust cross-reactivity to SARS-CoV-2 ^22,23^. However, the participants in this study did not show T cell responses against SARS-CoV-2 spike antigens on day 1, implying that none of the participants had pre-existing cross-reactive memory T cells against SARS-CoV-2.

The strengths of the present study include the longitudinal design with repeated measures of antibodies and T cell responses during and after the COVID-19 vaccination regimen. However, this study has several limitations. First, our study was conducted in a small-scale cohort of hospital workers in a single medical institution in Japan; thus, our findings might not be generalized to other settings. Second, the sufficient level of neutralizing activity or cellular immunities that is enough to protect against SARS-CoV-2 infection is not well known. In this study, sufficient neutralizing activity levels were defined as IH ≥35% in ELISA-based semi-quantitative neutralization assay (NeutraLISA), and the positive results evaluated by NeutraLISA spanned a wide range of values when the samples were analyzed by an *in vitro* virological experiment (summarized in Supplementary Figure S2). Further studies are warranted to determine sufficient levels of SARS-CoV-2 IgG antibody, neutralizing antibody, and T cell responses against SARS-CoV-2 infection and the duration of immunity induced by the vaccine.

In conclusion, our study demonstrated that BNT162b2 vaccination induces SARS-CoV-2 specific IgG-S antibody and T cell responses, which were detected on day 15 following the first dose and earlier than the onset of neutralizing activity, peaked after the second dose and gradually declined over time. Sufficient levels of these responses against SARS-CoV-2 infection were maintained for over six–ten weeks but not for seven months or later following the second dose of BNT162b2 vaccination, which may indicate the need for the booster dose. Our study contributes to a better understanding of the humoral and cellular immune responses upon COVID-19 vaccination.

## Data Availability

The data are not publicly available due to ethical restrictions for public deposition but available from the Center for Clinical Sciences, National Center for Global Health and Medicine, Tokyo, Japan (corresponding author: Wataru Sugiura) for researchers who meet the criteria to access the data.

## ACKNOWLEDGMENTS

We thank the members of the study group (Yusuke Oshiro, Natsumi Inamura, Takashi Nemoto, Haruka Osawa, Maki Konishi, and Nobumi Katayama) for their support. This work was funded by Abbott Japan (grant number 20C050), the NCGM COVID-19 Gift Fund (grant number 19K059), the Japan Health Research Promotion Bureau Research Fund (grant number 2020-B-09), AMED (grant number JP20nk0101627 and JP21nf0101627), and a grant from the National Center for Global Health and Medicine (grant number 21A006).

## AUTHOR CONTRIBUTIONS

WS, GU, and TM designed the study concept and methodology. AF, SY, and TM performed project administration and developed software. JST, AT, Kouki M, AK, and YK performed experiments. JST, AF, and SY analyzed and validated the data, and wrote the original draft manuscript. WS, NO, HM, MU, TM, MK, and Kenji M provided resources and contributed to supervision. All authors read and approved the final manuscript.

