## Supplementary Information for "SARS-CoV-2 specific T cell and humoral immune responses upon vaccination with BNT162b2: A 9 months longitudinal study"

Wataru Sugiura

Center for Clinical Sciences, National Center for Global Health and Medicine, 1-21-1, Toyama, Shinjuku-ku, Tokyo 162-8655, Japan

**This PDF file includes:**

Supplementary Table S1

Supplementary Figures S1 and S2

**Supplementary Table S1. Estimated geometric means with 95% confidence intervals of SARS-CoV-2 spike specific IgG titers by background factors**

| | | Day 1 (n=100) | | Day 15 (n=100) | | Day 29 (n=97) | | Day 61 (n=97) | | Day 82–96 (n=96) | | Day 224–232 (n=66) | | Day 263 (n=19) | | *P* for time | *P* for groups | *P* for interaction |
|---|---|---|---|---|---|---|---|---|---|---|---|---|---|---|---|---|---|---|
| | **N** | **GMT** | **95% CI** | **GMT** | **95% CI** | **GMT** | **95% CI** | **GMT** | **95% CI** | **GMT** | **95% CI** | **GMT** | **95% CI** | **GMT** | **95% CI** | | | |
| **Age <40** | 42 | 2.4 | 1.9–3.0 | 726 | 581–908* | 22,671 | 17,926–28,639* | 9,571 | 7,491–12,228 | 5,621 | 4,325–7,307* | 1,015 | 736–1,398* | 956 | 569–1,606 | <0.01 | <0.01 | 0.01 |
| **Age ≥40** | 58 | 2.4 | 2.0–2.9 | 400 | 331–484* | 15,239 | 12,558–18,492* | 7,025 | 5,736–8,602 | 3,963 | 3,188–4,928* | 610 | 470–793* | 501 | 323–777 | | | |
| **Men** | 32 | 2.1 | 1.7–2.8 | 427 | 330–552 | 14,935 | 11,488–19,417 | 6,804 | 5,169–8,956 | 3,965 | 2,948–5,334 | 713 | 500–1,016 | 561 | 307–1,022 | <0.01 | 0.14 | 0.96 |
| **Women** | 68 | 2.5 | 2.1–3.0 | 559 | 469–667 | 19,566 | 16,294–23,497 | 8,609 | 7,107–10,430 | 4,900 | 3,992–6,015 | 768 | 598–985 | 702 | 466–1,058 | | | |
| **BMI <25** | 83 | 2.4 | 2.1–2.8 | 531 | 453–624 | 18,490 | 15,682–21,800 | 8,114 | 6,829–9,641 | 4,727 | 3,932–5,682 | 774 | 619–967 | 657 | 450–960 | <0.01 | 0.41 | 0.99 |
| **BMI ≥25** | 17 | 2.3 | 1.6–3.2 | 428 | 299–612 | 15,320 | 10,540–22,269 | 7,323 | 4,951–10,829 | 3,865 | 2,525–5,916 | 644 | 389–1,066 | 612 | 286–1,308 | | | |

Data are shown as geometric means with 95% confidence intervals estimated by the repeated measures mixed model.

All models were adjusted for age (<40 or ≥40 years) and sex.

*Significant differences between groups at each time point (*P*<0.05).

GMT: geometric mean titers.

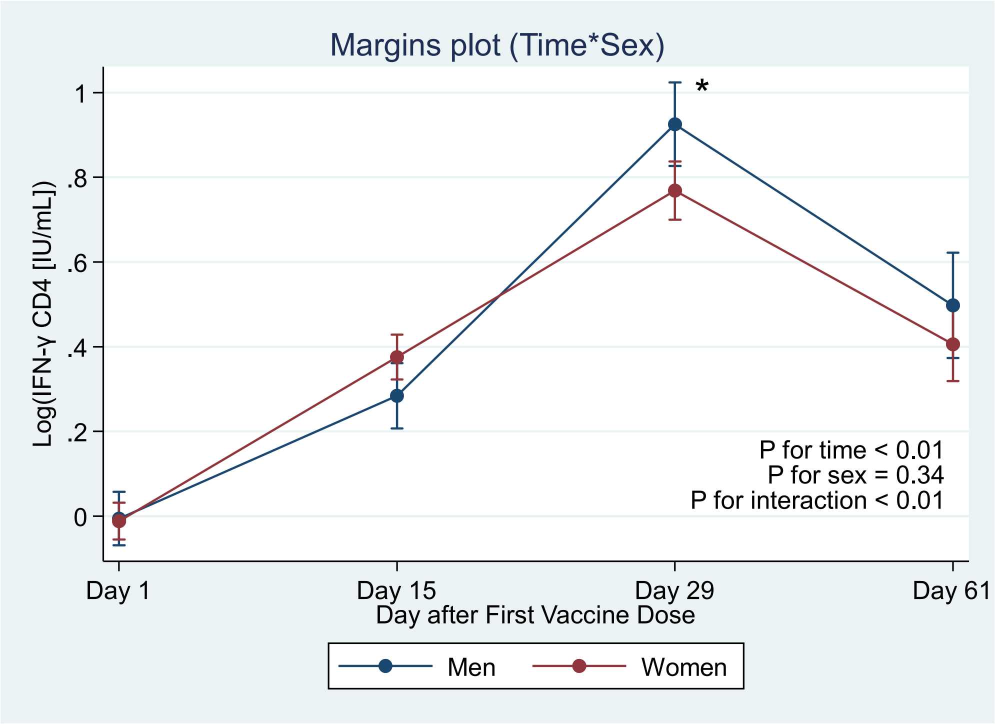


**Supplementary Figure S1. Estimated geometric means with 95% confidence intervals of IFN-γ $CD4^{+}$ T cell by background factor (sex)**

We fitted a mixed model of repeated measures using an unstructured covariance matrix. Background factors were treated as categorical variables in the models: sex (men or women). $Log_{10}$-transformed IFN-γ outcomes were used as dependent variables. We considered each background factor (sex), time, and interaction as fixed factors, and individual identifiers and time as random factors in the model. We estimated the mean $log_{10}$-transformed IgG-S titer with 95% confidence intervals (CIs); then, we back-transformed them and presented them as geometric means. To compare the mean differences of IFN-γ outcomes between background factors at each time point, we used the Wald test with Bonferroni adjustment. Blue and red lines indicate men and women, respectively.

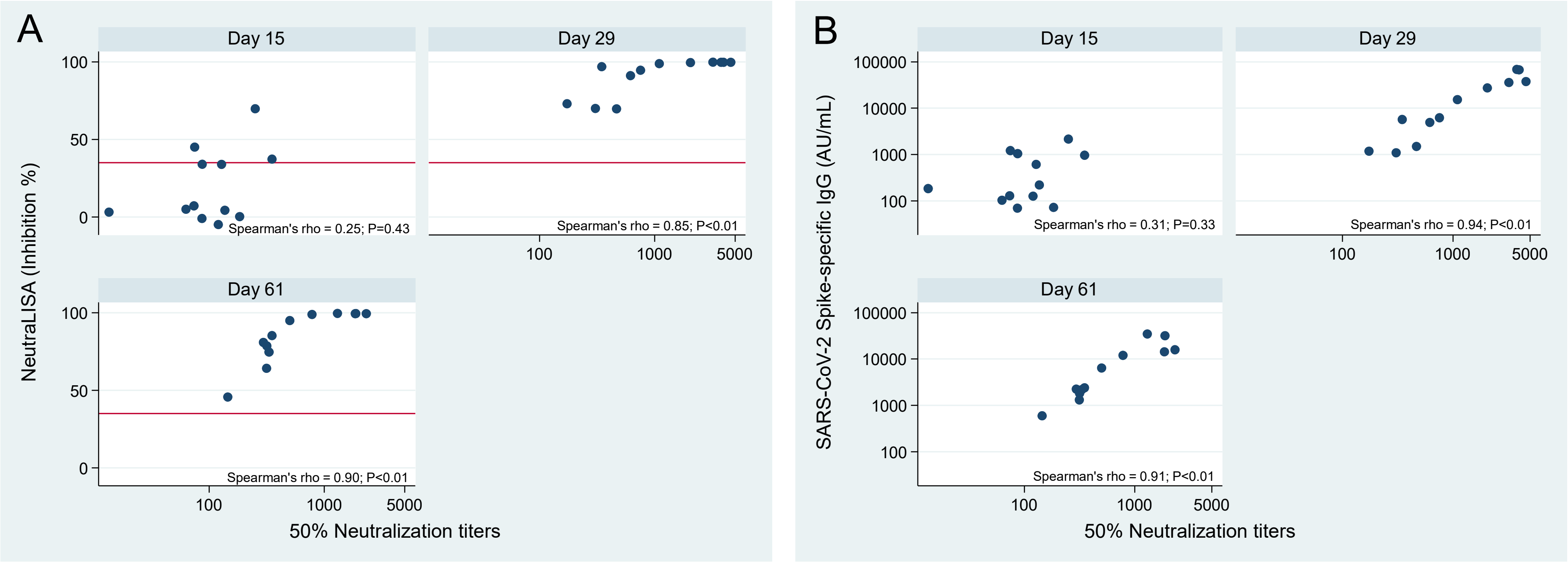


**Supplementary Figure S2. Scatter plot of 50% Neutralization titers with NeutraLISA (A) or IgG-S titers (B)**

To evaluate the results of the ELISA-based semi-quantitative neutralization assay (SARS-CoV-2-NeutraLISA kit, Euroimmun), an in vitro virological experiment-based neutralizing assay was performed using 36 serum samples (12 samples × 3 time points). Although all samples were evaluated as positive for neutralizing antibodies by NeutraLISA on days 29 and 61, the serum dilution that resulted in 50% neutralization titers ($NT_{50}$) spanned a wide range of values (174 to 4585-fold and 145 to 2312-fold dilution for days 29 and 61, respectively). Red lines indicate the cutoff (35 %) for the NeutraLISA assay. At day 15, neither NeutraLISA nor IgG-S titers were correlated with 50% neutralization titers. At days 29 and 61, NeutraLISA and IgG-S titers were significantly correlated with 50% neutralization titers, although the results of NeutraLISA appeared to be saturated at around 1000 titers of 50% neutralizing antibody.
